# Glucose metabolism and Radiodensity of Abdominal Adipose Tissue: A 5-year longitudinal study in a large PET cohort

**DOI:** 10.1101/2024.01.21.24301581

**Authors:** Kyoungjune Pak, Severi Santavirta, Seunghyeon Shin, Hyun-Yeol Nam, Sven De Maeyer, Lauri Nummenmaa

## Abstract

**Objective:** ^18^F-Fluorodeoxyglucose positron emission tomography/computed tomography (PET/CT) allows the noninvasive assessment of glucose metabolism and radiodensity of visceral adipose tissue (VAT) and subcutaneous adipose tissue (SAT).

**Research design and methods:** We retrospectively analyzed data from 435 healthy males (mean 42.8 years) who underwent health check-up program twice at baseline and 5-year follow-up. The mean standardized uptake value (SUV) was measured from SAT and VAT and was divided with liver SUV. The mean Hounsfield unit (HU) of SAT and VAT was measured from CT scans. The effects of clinical variable clusters on SUVR were investigated using Bayesian hierarchical modelling. Four clusters were established for predicting SUVR; 1) metabolic cluster (BMI, waist-to-hip ratio, fat percentage, muscle percentage*-1, HOMA-IR), 2) blood pressure (systolic, diastolic), 3) glucose (fasting plasma glucose level, HbA1c), and 4) C-reactive protein.

**Results:** All the clinical variables except for C-reactive protein changed during the 5-year follow-up. SUVR and HU of VAT were increased during the 5-year follow-up, however, those of SAT were not changed. SUVR and HU were positively correlated in both VAT and SAT. SAT SUVR and VAT SUVR were negatively associated with metabolic cluster.

**Conclusion:** Ageing led to increased glucose metabolism and radiodensity in VAT, not in SAT. VAT may reflect the ageing process more directly than SAT. Glucose metabolism was higher and radiodensity was lower in VAT than in SAT, probably due to the difference in gene expression and lipid-density. Both glucose metabolism and radiodensity of VAT and SAT reflect the metabolic status.

## INTRODUCTION

The prevalence of obesity has nearly tripled in the last three decades (1). Obesity has become one of the most critical public health concern in the 21^st^ century (1). It is associated with an increased risk of a series of chronic metabolic diseases, such as cardiovascular disease, diabetes, dyslipidemia, hypertension, and non-alcoholic fatty liver (2; 3).

Adipose tissue was for a long time regarded as a tissue without a specific anatomy (4). It consists of multiple depots situated in two primary compartments within the body: visceral adipose tissue (VAT) and subcutaneous adipose tissue (SAT). VAT is located within the intra-abdominal cavity and is linked to digestive organs, includes omental, mesenteric and epiploic adipose tissue depots (4; 5). SAT is located beneath the skin, separated by abdominal wall, representing over 80% of total body fat (5). Accumulating evidence however suggests that adipose depots are structured to form a large organ with distinct anatomy, specific vascular and nerve supplies, complex cytology and high physiological plasticity (6). In addition, numerous studies have demonstrated the differences between VAT and SAT in terms of gene expression profiles, cell morphology and the association with the cardiometabolic diseases (6). Therefore, it is possible that inflammation within each of VAT and SAT could carry varying implications for the risk of cardiometabolic diseases (7).

^18^F-Fluorodeoxyglucose (FDG) positron emission tomography (PET) allows the noninvasive assessment of glucose metabolic activity and inflammation in the adipose tissue (8). Also, computed tomography (CT) enables a noninvasive evaluation of adipose tissue quality in Hounsfield unit (HU) (9). Thus, ^18^F-FDG PET/CT is a reliable tool to evaluate VAT and SAT with these two modalities at once. Glucose metabolism of VAT is lower in obese subjects than lean subjects (10) and the lower attenuation of VAT is associated with risk factors of cardiometabolic diseases (11). However, there has been no investigation on the interaction between glucose metabolism and radiodensity of abdominal adipose tissue, the distinctive features of VAT and SAT in terms of glucose metabolism and radiodensity and the longitudinal changes of VAT and SAT over time. Therefore, to address the effects of ageing and metabolic factors on abdominal adipose tissue, we analyzed a large cohort (n=435) of healthy middle-aged adults (mean 42.8 years) who underwent ^18^F-FDG PET/CT scans and a health check-up program twice: at the baseline and at 5-year follow-up. We used Bayesian hierarchical modeling to estimate the effects of clinical variables on glucose metabolism and radiodensity of VAT and SAT.

## RESEARCH DESIGN AND METHODS

### Subjects

We retrospectively analyzed data from 473 healthy males who underwent health check-up program at Samsung Changwon Hospital Health Promotion Center in 2013 (baseline) and 2018 (follow-up). After excluding subjects with neuropsychiatric disorders (n=5) or malignancies (n=3), those with missing data of anthropometric and body composition measurements (n=24) or CT (n=6), 435 healthy males were included in both baseline (mean 42.8, range 38-50 years) and follow-up (mean 48.0, range 43-55 years) studies. Health check-up program included 1) ^18^F-FDG PET/CT, 2) anthropometric and body composition measurements and 3) blood samples. Subjects in this study were included in a previous study of the effect of ageing on brain glucose metabolism (12). The study protocol was approved by the Institutional Review Board (Samsung Changwon Medical Ccenter 2019-06-005-004) and the informed consent from the participants was waived due to the retrospective study design.

### 18F-FDG PET/CT

Subjects were asked to avoid strenuous exercise for 24 hours and fast for at least 6 hours before PET study. PET/CT was performed 60 mins after injection of ^18^F-FDG (3.7 MBq/kg) with Discovery 710 PET/CT scanner (GE Healthcare, Waukesha, WI, USA). Continuous spiral CT was obtained with a tube voltage of 120kVp and tube current of 30-180mAs. PET scan was obtained in 3-dimensional mode with full width at half maximum of 5.6 mm and reconstructed using an ordered-subset expectation maximization algorithm. From PET scans, the mean standardized uptake value (SUV) was measured with the circular region-of-interest (ROI) from right lobe of the liver parenchyma, SAT and VAT at umbilicus level. The standardized uptake value ratio (SUVR) was calculated after dividing SUV from each ROI with liver SUV. Using the same ROIs with SUV measurement, the mean Hounsfield unit (HU) of SAT and VAT was measured from CT scans. A representative PET/CT scan with ROI is shown in Figure 1.

**Figure 1.**
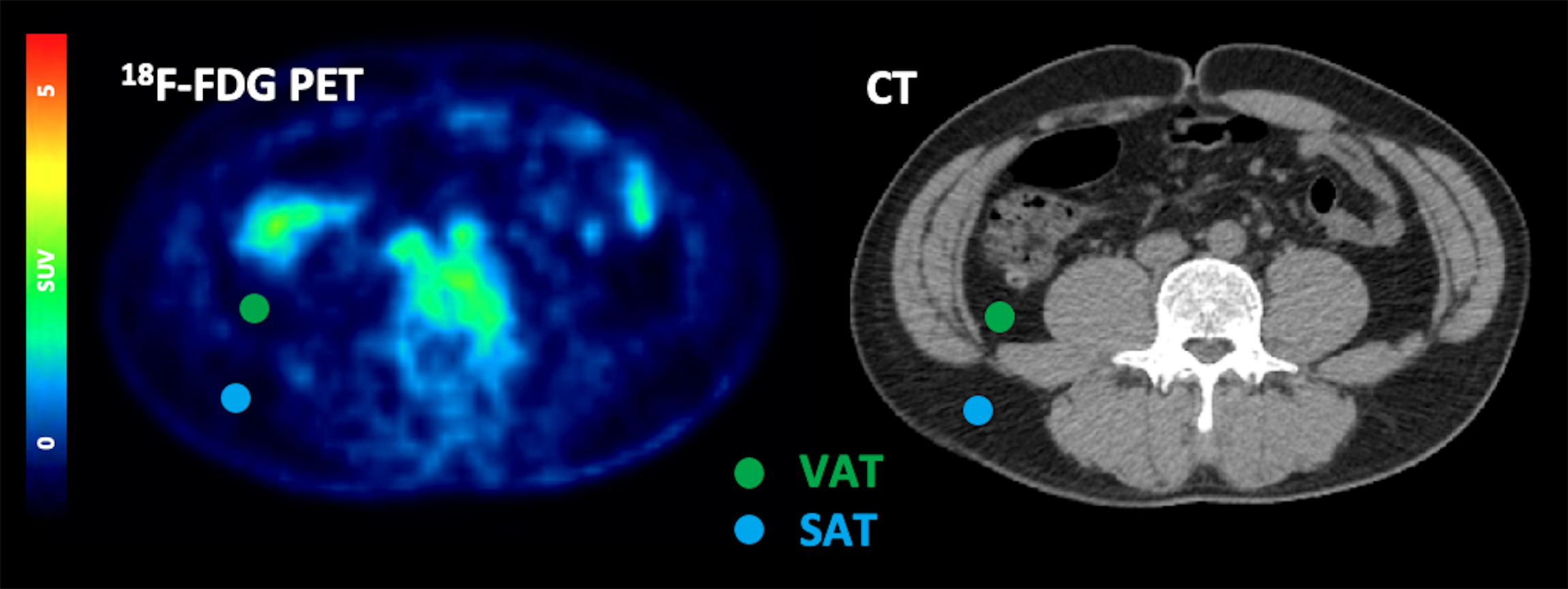
A representative PET/CT scan with ROI.

### Body measurement

For anthropometric measurements, height (cm) and weight (kg) were measured and body mass index (BMI) was calculated as weight/height^2^ (kg/m^2^). Waist and hip circumference (cm) were measured, and wait-to-hip ratio was calculated. For body composition measurement, a commercially available bioelectrical impedance analysis (InBody S10, Biospace, Seoul, Republic of Korea) was used to calculate fat percentage (%) and muscle percentage (%) after dividing fat mass (kg) and skeletal muscle mass (kg) with weight (kg). Systolic and diastolic blood pressures (mmHg) and heart rate were measured using an automatic sphygmomanometer (EASY X 800; Jawon Medical Co., Ltd, Seoul, Korea) after at least 10 mins of rest.

### Blood samples

Blood samples were collected from the antecubital vein of each subject. Fasting plasma glucose (mg/dL) and insulin (μIU/mL) were measured and homeostatic model assessment for insulin resistance (HOMA-IR) was calculated as follows: fasting insulin (μIU/mL)*fasting plasma glucose (mg/dL)/405 (13). Hemoglobin A1c (HbA1c, %) levels were analyzed with high-performance liquid chromatography.

### Statistical analysis

Normality was tested with Shapiro-Wilk test and SUVR and HU were log-transformed after adding 130 to HU as the minimum HU was -129 (baseline VAT). Comparison of clinical variables and SUVRs between baseline and follow-up was assessed with paired t-test. Pearson correlation was used to determine the association of SUVR and HU.

### Cluster analysis for predictor variables

Because clinical variables contained missing values (baseline study: HOMA-IR 35/435), we used a non-parametric imputation algorithm missForest with default parameters for imputation of the missing values (14). As some clinical variables were correlated, we used hierarchical clustering analysis before the Bayesian hierarchical modelling (see ref (12)). Before clustering, muscle percentage was multiplied with -1 to simplify the solution as it was the only variable showing negative correlations with the other predictors. Clustering yielded stable cluster hierarchy across the 6 tested algorithms (complete-linkage, single-linkage, UPGMA, WPGMA, WPGMC and Ward) and the following clusters were defined 1) metabolic cluster (BMI, waist-to-hip ratio, fat percentage, muscle percentage*-1, HOMA-IR), 2) blood pressure (systolic, diastolic), 3) glucose (fasting plasma glucose, HbA1c), and 4) C-reactive protein (CRP). In each cluster, the value was calculated after averaging the standardization of clinical variables (Supplementary Figure 1).

### Bayesian hierarchical modelling

The effects of clinical clusters on SUVR and HU were investigated using Bayesian hierarchical modelling with brms (15–17) that applies the Markov-Chain Monte Carlo sampling tools of RStan (18). We set up a separate model with SUVR or HU as a dependent variable and 4 clusters (metabolic, blood pressure, glucose, and CRP) as predictors. These fixed effects were calculated individually and as an interaction with time. We added subject and ROI as random intercepts to allow SUVR or HU to vary between subjects and ROIs and calculated the fixed effects including the interactions with time separately for each ROI as a random slope. Bayesian models were estimated using four Markov chains, each of which had 4,000 iterations including 1,000 warm-ups, thus totaling 12,000 post-warmup samples. The sampling parameters were slightly modified to facilitate convergence (max treedepth=20). Statistical analysis was carried out in R Statistical Software (The R Foundation for Statistical Computing).

## RESULTS

Subject characteristics are summarized in Table 1. Except for CRP (p=0.2450), all the clinical variables were increased during the 5-year follow-up; BMI (p=0.0002), waist-hip ratio (p<0.0001), fat percentage (p=0.0022), muscle percentage*-1 (p=0.0002), HOMA-IR (p<0.0001), systolic blood pressure (p<0.0001), diastolic blood pressure (p<0.0001), fasting plasma glucose (p<0.0001), and HbA1c (p<0.0001) (Supplementary Figure 2). VAT SUVR (p<0.0001) and VAT HU (p=0.0094) increased during the 5-year follow-up, however, SAT SUVR (p=0.2181) and SAT HU (p=0.2697) did not change significantly (Figure 2). SUVR and HU were positively correlated in baseline (SAT, r=0.5258, p<0.0001; VAT, r=0.4693, p<0.0001) and follow-up (SAT, r=0.5664, p<0.0001; VAT, r=0.4102, p<0.0001) studies (Figure 3). VAT SUVR was higher than SAT SUVR, while VAT HU was lower than SAT HU in both baseline and follow-up studies (all ps<0.0001). The effects of clinical variables on SUV and HUs were generally similar in the baseline and follow-up studies. SAT SUVR and VAT SUVR were negatively associated with metabolic cluster. SAT SUVR was positively associated with blood pressure and glucose. The effects of CRP markedly overlapped with zero in the analysis of both SAT and VAT (Figure 4). Both SAT HU and VAT HU were negatively associated with metabolic cluster. However, the effects of blood pressure, glucose, and CRP markedly overlapped with zero in both SAT and VAT (Figure 5).

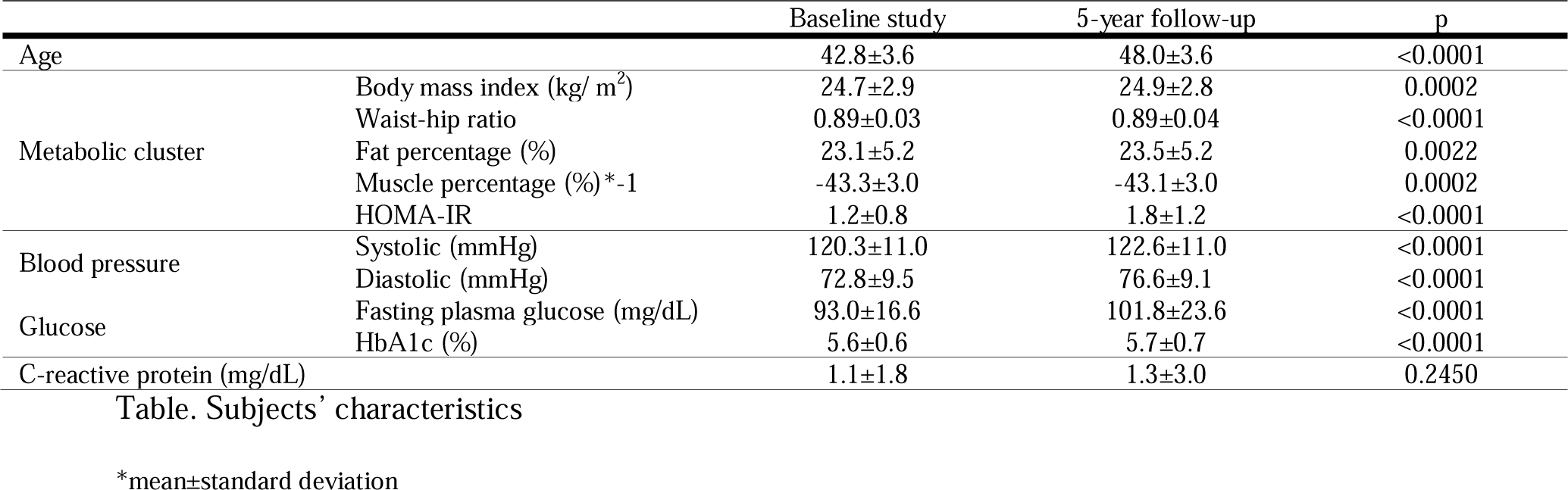

**Figure 2.**
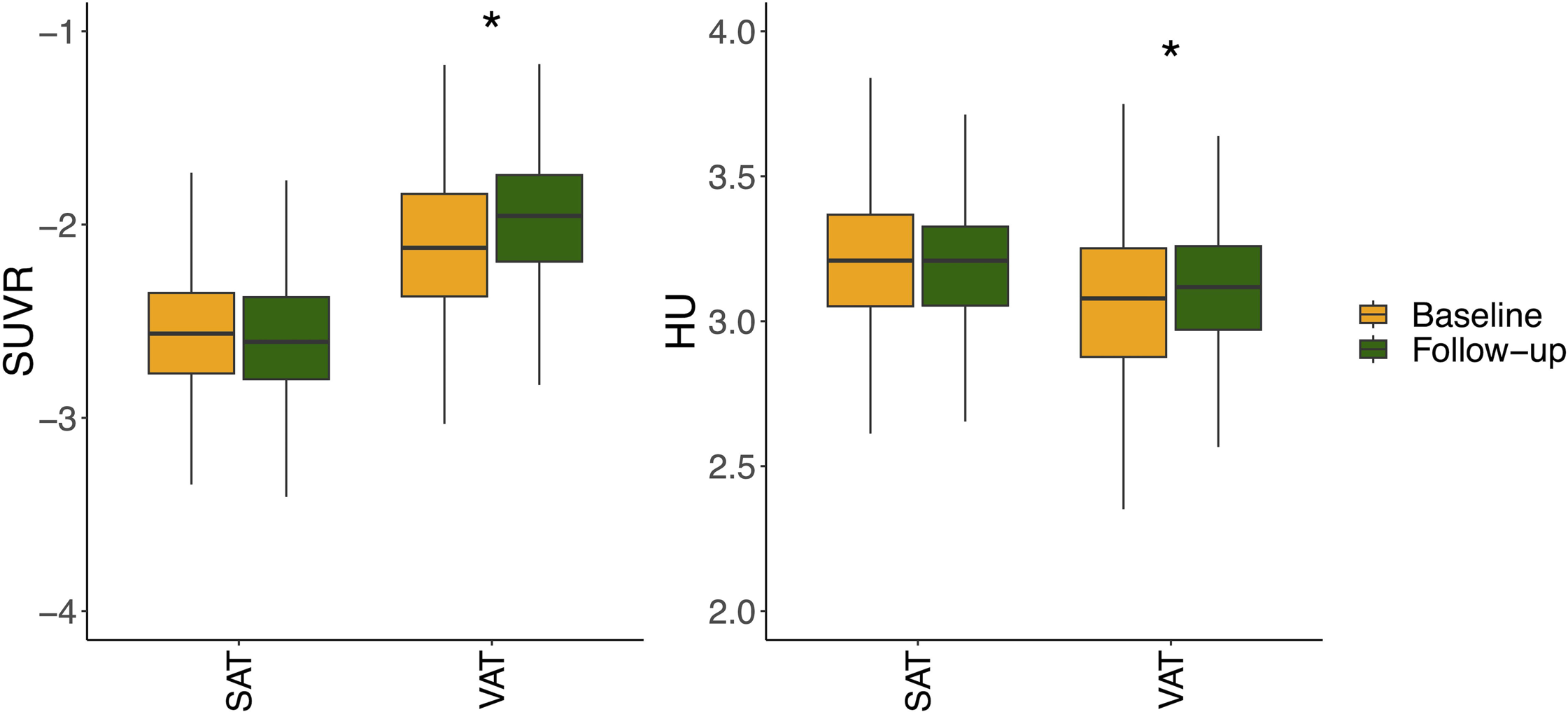
Comparison between baseline and follow-up studies: VAT SUVR (p<0.0001) and VAT HU (p=0.0094) increased during the 5-year follow-up, however, SAT SUVR (p=0.2181) and SAT HU (p=0.2697) did not change significantly.

**Figure 3.**
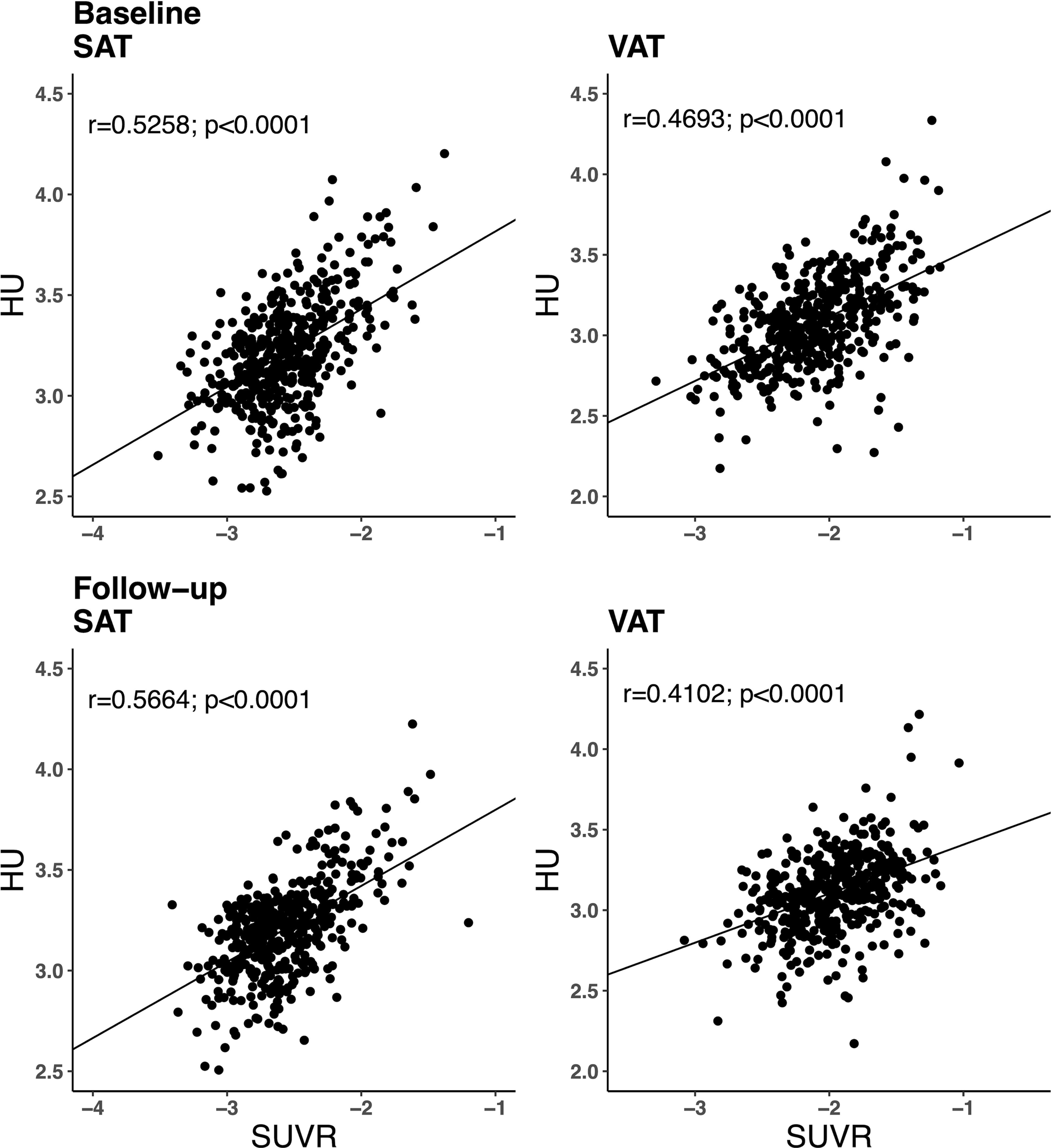
Correlation between SUVR and HU of SAT and VAT in baseline and follow-up studies: SUVR and HU were positively correlated in baseline (SAT, r=0.5258, p<0.0001; VAT, r=0.4693, p<0.0001) and follow-up (SAT, r=0.5664, p<0.0001; VAT, r=0.4102, p<0.0001) studies.

**Figure 4.**
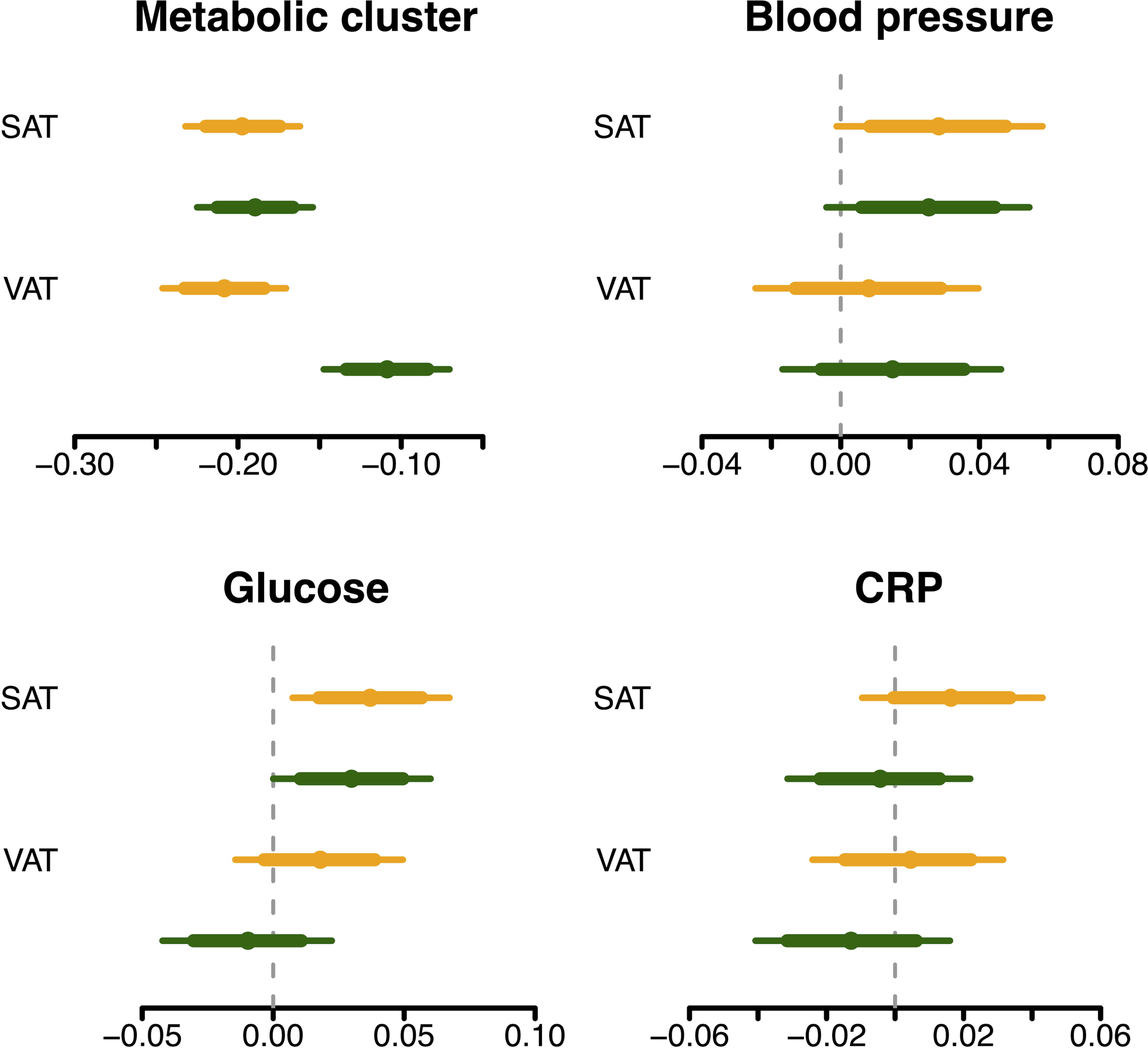
Posterior intervals of the regression coefficients for each cluster predicting SUVR. The thick lines represent the 80% posterior intervals, the thin lines represent the 95% posterior intervals, and the circles represent posterior means

**Figure 5.**
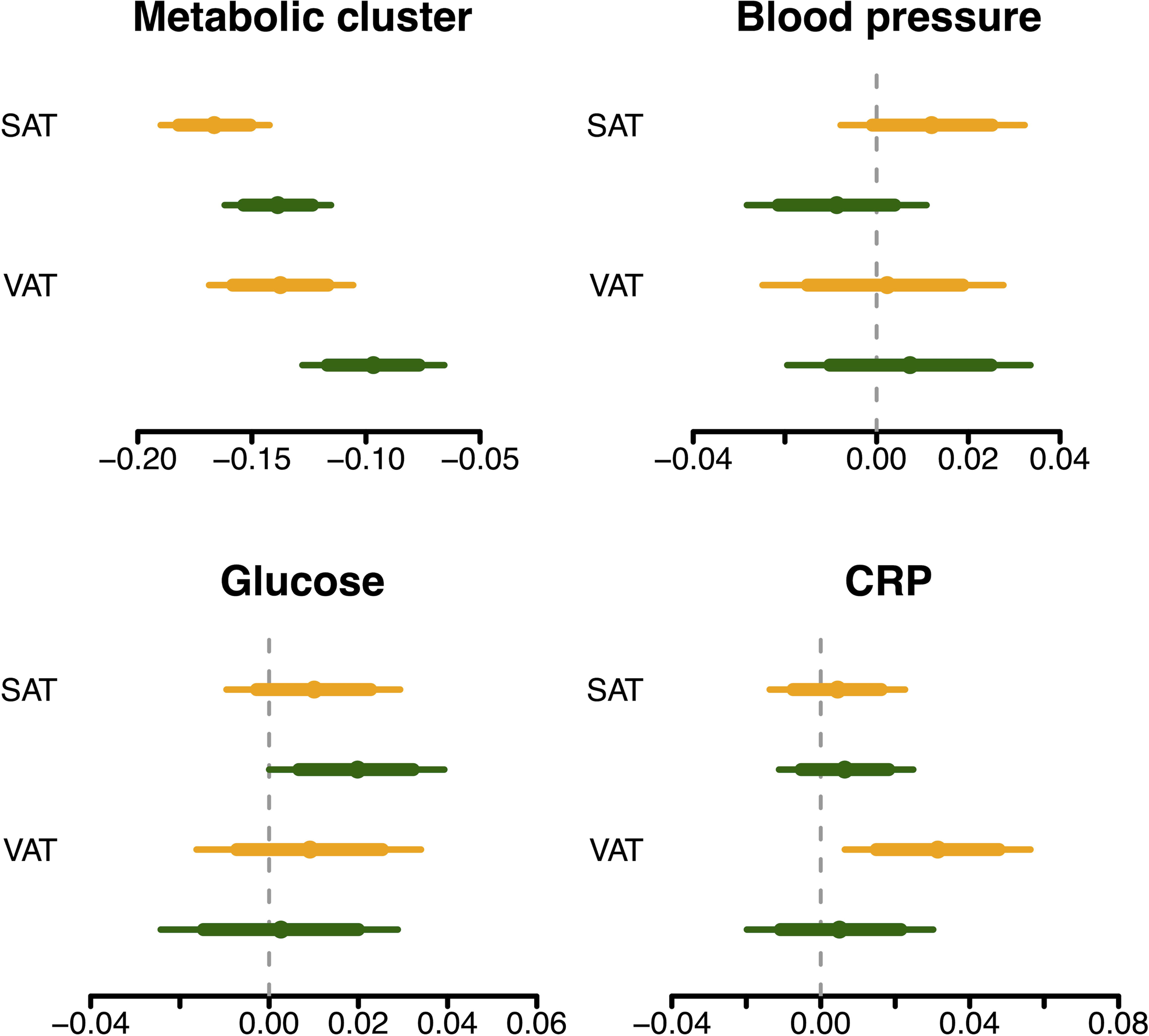
Posterior intervals of the regression coefficients for each cluster predicting HU. The thick lines represent the 80% posterior intervals, the thin lines represent the 95% posterior intervals, and the circles represent posterior means

## DISCUSSION

Our main finding was that ageing led to increased glucose metabolism and radiodensity in VAT, not in SAT. In both VAT and SAT, glucose metabolism and radiodensity were positively correlated each other. In VAT, glucose metabolism was higher and radiodensity was lower than SAT. Both glucose metabolism and radiodensity of VAT and SAT were negatively associated with metabolic cluster. Both glucose metabolism and radiodensity of SAT were positively associated with glucose. These effects of clinical variables on glucose metabolism and radiodensity were generally similar in the baseline and follow-up studies.

### VAT and SAT

VAT and SAT constitute two primary fat compartments linked to a number of cardiometabolic diseases (19). However, they have distinct functions and characteristics (5). VAT is located within the intra-abdominal cavity and it can engage in dynamic communication with neighboring internal organs (5). SAT in turn is located beneath the skin, separated by abdominal wall, providing protection against physical damage (5). VAT tends to exhibit pro-inflammatory characteristics. A certain portion of visceral adipogenic progenitors (APs) cells lose adipogenic potential and are transformed into pro-inflammatory fibrocytes, which exacerbates adipose tissue inflammation via inducing fibrosis and secreting pro-inflammatory cytokines such as tumor necrosis factor-alpha, interleukin-6, and monocyte chemotactic protein-1 (20). On the other hand, SAT has lower susceptibility to inflammatory responses in obesity. Subcutaneous adipogenic progenitors inhibit the infiltration of pro-inflammatory macrophage into SAT in response to gamma-aminobutyric acid, playing a crucial role in suppressing inflammation of SAT in obesity (21).

^18^F-FDG PET allows the noninvasive assessment of glucose metabolic activity and inflammation in the adipose tissues (8). Also, CT enables a noninvasive assessment of fat quality in HU (9). Thus, ^18^F-FDG PET/CT is a reliable tool for the evaluation of VAT and SAT with these two modalities at once. Prior studies have shown that VAT SUVR is lower in obese subjects than in lean subjects (10) and lower VAT HU was associated with increased risk of hypertension, insulin resistance and metabolic syndrome (11). On the other hand, SAT SUVR is not different between obese and lean subjects (10) and lower SAT HU is associated with decreased risk of diabetes and cardiometabolic risk factors (11). In the present study, VAT SUVR was higher than SAT SUVR, while VAT HU was lower than SAT HU. As FDG is an analogue of glucose, the significantly higher expression of hexokinase-1 gene in VAT might be the underlying mechanism of this difference of glucose metabolism between VAT and SAT (8). Lower HU in VAT represents more lipid-dense fat tissue (22). Adipose tissue with higher HU has been associated with smaller adipocytes probably due to extracellular matrix of adipose tissue (23). Larger adipocyte size may indicate an inability to generate new adipocyte cells and is linked with insulin resistance (24). Also, lower VAT HU may indicate relative lack of vascularity, as blood has higher HU than adipose tissue on CT (25). Therefore, VAT might be a tissue with higher glucose metabolism, more lipid-density and less vascularity than SAT.

### Ageing effect

Physiological changes occur with ageing in the brain and also in the body organs. Adiposity develops between the third and seventh decades of life and increased VAT accumulation is strongly linked to ectopic fat deposition in skeletal muscle, heart, liver, pancreas, or blood arteries (26). Also, the heart and blood vessels become stiffer, causing the blood pressure to be increased (27). Impaired insulin sensitivity and the increased risk of insulin resistance leads to the increase of fasting plasma glucose and HbA1c (28). In this study, all the clinical variables except for CRP increased during the 5-year follow-up: BMI, waist-hip ratio, fat percentage, muscle percentage*-1, HOMA-IR, blood pressure, fasting plasma glucose and HbA1c, similar with previous longitudinal studies (29). This shows the ageing process as a transition towards the loss of metabolic homeostasis and plasticity in the middle adulthood. Ageing also led to the increase of VAT SUVR and VAT HU, without the change in SAT SUVR and SAT HU. Therefore, VAT may reflect the ageing process more directly than SAT as SAT did not show the significant changes of ageing process in this study.

### Glucose metabolism, Radiodensity and Clinical variables

Inflammatory cells demonstrate the elevated affinity of glucose transporters for deoxyglucose as well as elevated expression of glucose transporters (30). Therefore, ^18^F-FDG PET is utilized for the diagnosis of infection and inflammation in clinical setting (30). However, different from our expectation, both VAT SUVR and SAT SUVR were negatively associated with metabolic cluster. Insulin-resistant adipocyte might overweigh FDG uptake from the inflammation of VAT and SAT (8). Also, lower SUVR may indicate abnormal perfusion, vascular function, and capillary density of VAT and SAT in higher metabolic cluster (31). SUVR and HU were positively correlated with each other and the effects of clinical variables were generally similar on both of SUVR and HU. Therefore, both SUVR and HU of VAT and SAT strongly reflect the metabolic status of the body. In addition to metabolic cluster, glucose cluster was positively associated with SUVR and HU of SAT, not with those of VAT. This is unexpected, as the effect of glucose cluster shows the opposite direction with that of metabolic cluster. Until now, the impact of glucose level on FDG uptake was investigated in the body organs such as lung, bone marrow, spleen, bowel and stomach (32), not in abdominal adipose tissue. FDG uptake of adipose tissue from upper back showed the positive correlation with glucose level (33), however, a study of Büsing et al (34) showed a neglectable change of FDG uptake in adipose tissue (area not specified) that was not influenced by glucose level.

### Limitations

Only males were included in this study, thus these results may not directly generalize to females. In addition, this retrospective study was based on health check-up program. Therefore, the mechanisms underlying these findings are not fully clarified in this study.

In conclusion, ageing led to increased glucose metabolism and radiodensity in VAT, not in SAT. VAT may reflect the ageing process more directly than SAT. Glucose metabolism was higher and radiodensity was lower in VAT than in SAT, probably due to the difference in gene expression and lipid-density. Both glucose metabolism and radiodensity of VAT and SAT reflect the metabolic status of the body.

## Data Availability

Data sets generated during the current study are available from the corresponding author on reasonable request

## AUTHOR CONTRIBUTION

KP, TM, SS, LN researched data, contributed to discussion, and wrote, reviewed, and edited the manuscript. SS, HYN, SDM contributed to discussion and reviewed and edited the manuscript. All authors approved the final version of the manuscript. KP is the guarantor of this work and, as such, had full access to all the data in the study and takes responsibility for the integrity of the data and the accuracy of the data analysis.

## PRIOR PRESENTATION

A non–peer-reviewed version of this article was submitted to medRxiv preprint server on 17th January 2024.

## REFERENCES

1. Agha M, Agha R. The rising prevalence of obesity: part A: impact on public health. Int J Surg Oncol (N Y) 2017;2:e17

2. Eisenstein SA, Bischoff AN, Gredysa DM, Antenor-Dorsey JA, Koller JM, Al-Lozi A, Pepino MY, Klein S, Perlmutter JS, Moerlein SM, Black KJ, Hershey T. Emotional Eating Phenotype is Associated with Central Dopamine D2 Receptor Binding Independent of Body Mass Index. Sci Rep 2015;5:11283

3. Flegal KM, Carroll MD, Kuczmarski RJ, Johnson CL. Overweight and obesity in the United States: prevalence and trends, 1960-1994. International journal of obesity and related metabolic disorders : journal of the International Association for the Study of Obesity 1998;22:39–47

4. Passaro A, Miselli MA, Sanz JM, Dalla Nora E, Morieri ML, Colonna R, Pisot R, Zuliani G. Gene expression regional differences in human subcutaneous adipose tissue. BMC Genomics 2017;18:202

5. Hwang I, Kim JB. Two Faces of White Adipose Tissue with Heterogeneous Adipogenic Progenitors. Diabetes Metab J 2019;43:752–762

6. Cinti S. The adipose organ at a glance. Dis Model Mech 2012;5:588–594

7. Tahara N, Yamagishi S, Kodama N, Tahara A, Honda A, Nitta Y, Igata S, Matsui T, Takeuchi M, Kaida H, Kurata S, Abe T, Fukumoto Y. Clinical and biochemical factors associated with area and metabolic activity in the visceral and subcutaneous adipose tissues by FDG-PET/CT. J Clin Endocrinol Metab 2015;100:E739–747

8. Christen T, Sheikine Y, Rocha VZ, Hurwitz S, Goldfine AB, Di Carli M, Libby P. Increased glucose uptake in visceral versus subcutaneous adipose tissue revealed by PET imaging. JACC Cardiovasc Imaging 2010;3:843–851

9. Rosenquist KJ, Massaro JM, Pedley A, Long MT, Kreger BE, Vasan RS, Murabito JM, Hoffmann U, Fox CS. Fat quality and incident cardiovascular disease, all-cause mortality, and cancer mortality. J Clin Endocrinol Metab 2015;100:227–234

10. Oliveira AL, Azevedo DC, Bredella MA, Stanley TL, Torriani M. Visceral and subcutaneous adipose tissue FDG uptake by PET/CT in metabolically healthy obese subjects. Obesity (Silver Spring) 2015;23:286–289

11. Rosenquist KJ, Pedley A, Massaro JM, Therkelsen KE, Murabito JM, Hoffmann U, Fox CS. Visceral and subcutaneous fat quality and cardiometabolic risk. JACC Cardiovasc Imaging 2013;6:762–771

12. Pak K, Malen T, Santavirta S, Shin S, Nam HY, De Maeyer S, Nummenmaa L. Brain Glucose Metabolism and Aging: A 5-Year Longitudinal Study in a Large Positron Emission Tomography Cohort. Diabetes Care 2023;46:e64–e66

13. Matthews DR, Hosker JP, Rudenski AS, Naylor BA, Treacher DF, Turner RC. Homeostasis model assessment: insulin resistance and beta-cell function from fasting plasma glucose and insulin concentrations in man. Diabetologia 1985;28:412–419

14. Stekhoven DJ, Buhlmann P. MissForest--non-parametric missing value imputation for mixed-type data. Bioinformatics 2012;28:112–118

15. Bürkner P-C. Bayesian Item Response Modeling in R with brms and Stan. Journal of Statistical Software 2021;100:1–54

16. Bürkner P-C. brms: an R package for Bayesian multilevel models using Stan. Journal of Statistical Software 2017;80:1–28

17. Bürkner P-C. Advanced Bayesian multilevel modeling with the R package brms. The R Journal 2018;10:395–411

18. RStan: the R interface to Stan [article online], 2022. Available from https://mc-stan.org/.

19. Fox CS, Massaro JM, Hoffmann U, Pou KM, Maurovich-Horvat P, Liu CY, Vasan RS, Murabito JM, Meigs JB, Cupples LA, D’Agostino RB, Sr., O’Donnell CJ. Abdominal visceral and subcutaneous adipose tissue compartments: association with metabolic risk factors in the Framingham Heart Study. Circulation 2007;116:39–48

20. Weisberg SP, McCann D, Desai M, Rosenbaum M, Leibel RL, Ferrante AW, Jr. Obesity is associated with macrophage accumulation in adipose tissue. J Clin Invest 2003;112:1796–1808

21. Hwang I, Jo K, Shin KC, Kim JI, Ji Y, Park YJ, Park J, Jeon YG, Ka S, Suk S, Noh HL, Choe SS, Alfadda AA, Kim JK, Kim S, Kim JB. GABA-stimulated adipose-derived stem cells suppress subcutaneous adipose inflammation in obesity. Proc Natl Acad Sci U S A 2019;116:11936–11945

22. Wronska A, Kmiec Z. Structural and biochemical characteristics of various white adipose tissue depots. Acta Physiol (Oxf) 2012;205:194–208

23. Murphy RA, Register TC, Shively CA, Carr JJ, Ge Y, Heilbrun ME, Cummings SR, Koster A, Nevitt MC, Satterfield S, Tylvasky FA, Strotmeyer ES, Newman AB, Simonsick EM, Scherzinger A, Goodpaster BH, Launer LJ, Eiriksdottir G, Sigurdsson S, Sigurdsson G, Gudnason V, Lang TF, Kritchevsky SB, Harris TB. Adipose tissue density, a novel biomarker predicting mortality risk in older adults. J Gerontol A Biol Sci Med Sci 2014;69:109–117

24. Weyer C, Foley JE, Bogardus C, Tataranni PA, Pratley RE. Enlarged subcutaneous abdominal adipocyte size, but not obesity itself, predicts type II diabetes independent of insulin resistance. Diabetologia 2000;43:1498–1506

25. Pasarica M, Sereda OR, Redman LM, Albarado DC, Hymel DT, Roan LE, Rood JC, Burk DH, Smith SR. Reduced adipose tissue oxygenation in human obesity: evidence for rarefaction, macrophage chemotaxis, and inflammation without an angiogenic response. Diabetes 2009;58:718–725

26. Kuk JL, Saunders TJ, Davidson LE, Ross R. Age-related changes in total and regional fat distribution. Ageing Res Rev 2009;8:339–348

27. Buford TW. Hypertension and aging. Ageing Res Rev 2016;26:96–111

28. Shou J, Chen PJ, Xiao WH. Mechanism of increased risk of insulin resistance in aging skeletal muscle. Diabetol Metab Syndr 2020;12:14

29. Lee JJ, Pedley A, Hoffmann U, Massaro JM, Fox CS. Association of Changes in Abdominal Fat Quantity and Quality With Incident Cardiovascular Disease Risk Factors. J Am Coll Cardiol 2016;68:1509–1521

30. Love C, Tomas MB, Tronco GG, Palestro CJ. FDG PET of infection and inflammation. Radiographics 2005;25:1357–1368

31. Gealekman O, Guseva N, Hartigan C, Apotheker S, Gorgoglione M, Gurav K, Tran KV, Straubhaar J, Nicoloro S, Czech MP, Thompson M, Perugini RA, Corvera S. Depot-specific differences and insufficient subcutaneous adipose tissue angiogenesis in human obesity. Circulation 2011;123:186–194

32. Lindholm H, Brolin F, Jonsson C, Jacobsson H. The relation between the blood glucose level and the FDG uptake of tissues at normal PET examinations. EJNMMI Res 2013;3:50

33. Sharma P, Chatterjee P, Alvarado LA, Dwivedi AK. Standardized uptake value of normal organs on routine clinical [18F]FDG PET/CT: impact of tumor metabolism and patient-related factors. Nucl Med Rev Cent East Eur 2023;26:1–10

34. Busing KA, Schonberg SO, Brade J, Wasser K. Impact of blood glucose, diabetes, insulin, and obesity on standardized uptake values in tumors and healthy organs on 18F-FDG PET/CT. Nucl Med Biol 2013;40:206–213

