## Supplementary figure 1, 2 for "Glucose metabolism and Radiodensity of Abdominal Adipose Tissue: A 5-year longitudinal study in a large PET cohort"

**Supplementary Figure 1.** Four clusters after hierarchical clustering analysis

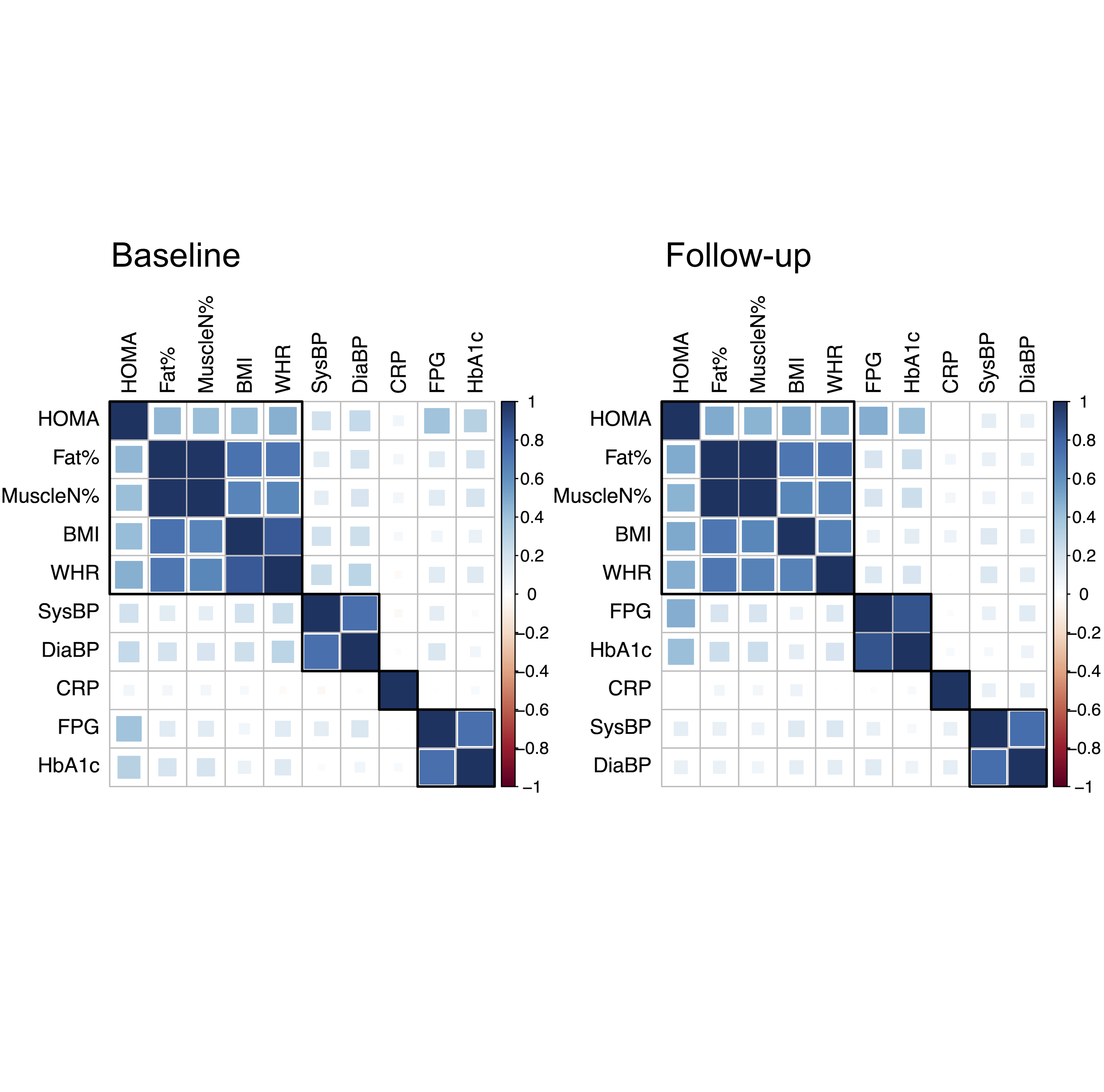

**Supplementary Figure 2.** All the clinical variables were increased during the 5-year follow-up except for CRP; BMI (p=0.0002), waist-hip ratio (p<0.0001), fat percentage (p=0.0022), muscle percentage*-1 (p=0.0002), HOMA-IR (p<0.0001), systolic blood pressure (p<0.0001), diastolic blood pressure (p<0.0001), fasting plasma glucose (p<0.0001), and HbA1c (p<0.0001).

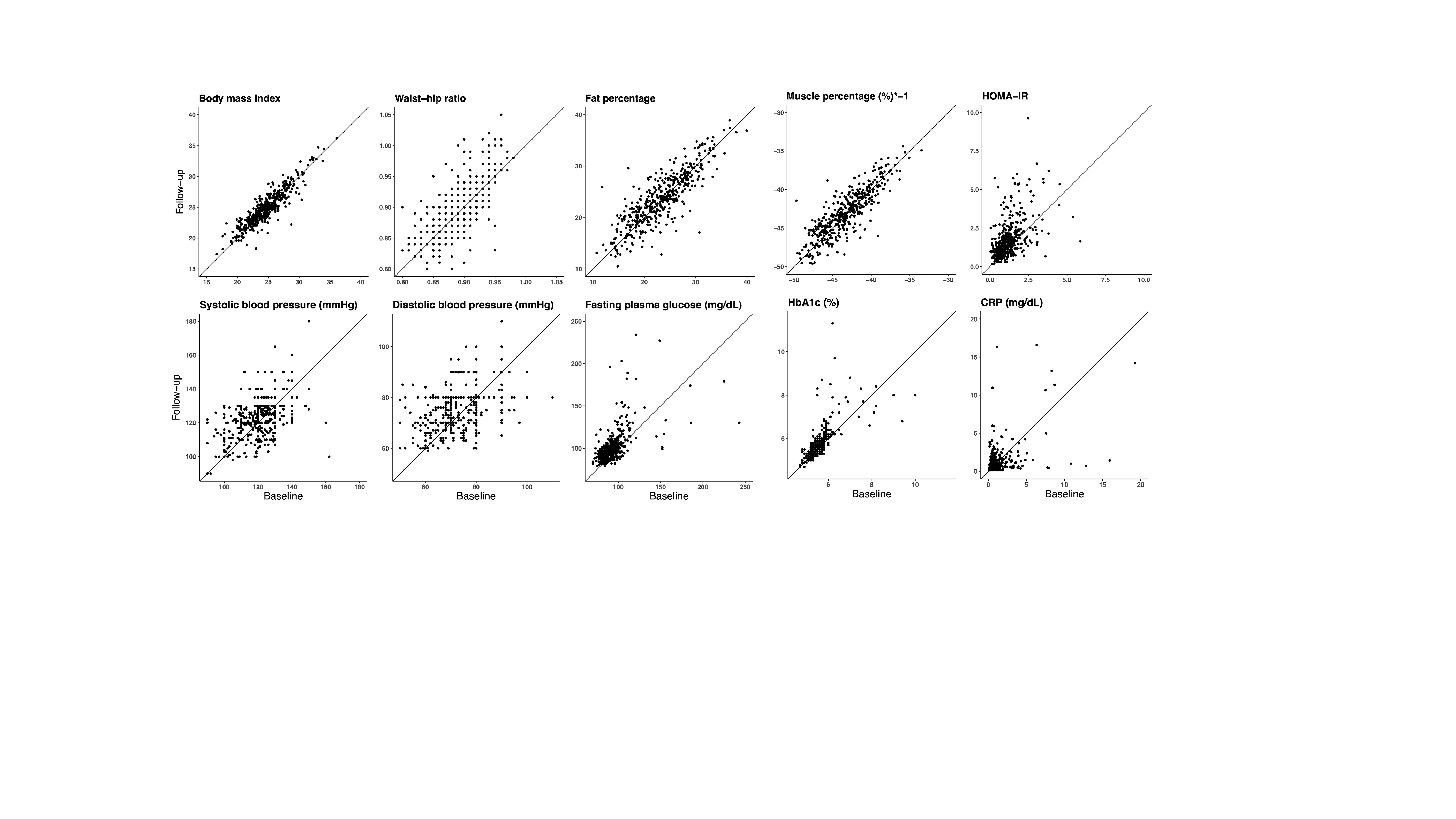
